# HIV PrEP programmes as a framework for diagnosing and treating HBV infection in adolescents and young adults in KwaZulu-Natal, South Africa

**DOI:** 10.1101/2024.11.29.24318178

**Authors:** Gloria Sukali, Jacob Busang, Jaco Dreyer, Thandeka Khoza, Marion Delphin, Nonhlanhla Okesola, Carina Herbst, Elizabeth Waddilove, Janine Upton, Janet Seeley, Collins Iwuji, Motswedi Anderson, Philippa C Matthews, Maryam Shahmanesh

**Affiliations:** Africa Health Research Institute, KwaZulu-Natal, South Africa; Division of Infection and Immunity, University College London, Gower Street, London, WC1E 6BT, UK; The Francis Crick Institute, 1 Midland Road, London, NW1 1AT, UK; Department of Global Health & Infection, Brighton and Sussex Medical School, University of Sussex, Falmer, Brighton, BN1 9PX, UK; Department of Infectious Diseases, University College London Hospital, Euston Road, London NW1 2BU, UK

**Author notes:** joint senior/corresponding.

**Keywords:** HIV, HBV, hepatitis B, South Africa, infection, prevention, prevalence, diagnosis, decentralisation, treatment, PrEP

## Abstract

**Background:** Hepatitis B virus (HBV) is a neglected public health threat with poor community awareness and access to prevention, despite having a safe and effective vaccine. There are still gaps in diagnosis and treatment, particularly in the World Health Organization (WHO) African region. New WHO HBV guidelines, for the first time, include the use of dual therapy for HBV treatment (Tenofovir (TDF) and Emtricitabine or Lamivudine (XTC) due to challenges in accessing TDF monotherapy. TDF/XTC is also recommended as Pre-Exposure Prophylaxis (PrEP) in adolescents and adults at risk of Human Immunodeficiency Virus (HIV).HBV Screening, treatment and prevention need to be decentralized to improve access. We hypothesize that HBV programmes in African settings can use pre-existing HIV infrastructure, in particular building on PrEP programmes, for access to TDF.

**Methods:** At the Africa Health Research Institute (AHRI) in KwaZulu Natal, South Africa, the new Evaluation of Vukuzazi LiVEr disease - Hepatitis B (‘EVOLVE-HBV’, UCL ethics ref. 23221/001) research programme explored the PrEP uptake and retention cascade amongst adolescents and youth aged 15-30 year-olds living with HBV through decentralized sexual health /HIV services of the ‘*Thetha nami ngithethe nawe’* and the Long-acting HIV Pre-Exposure Prophylaxis (LAPIS) study (UKZN BREC ethics ref. 473/2019 and 3735/2021). Following point of care testing (POCT) for HBsAg, follow-up venous samples were taken for laboratory confirmation.

**Results:** Over the time reviewed (May 2021 - Sept 2024), 15,847 adolescents and young adults received a ‘needs assessment’ by peer navigators in the community, of whom 3481 (21.9%) were eligible for HIV prevention interventions and referred for clinical review. 3431 (98.6%) accepted HBV POCT as part of routine screening, of whom 21 (0.6%) tested positive for HBsAg. These 21 individuals had not previously been aware of their HBV status, but one was already on antiretroviral (ART) for HIV infection. Amongst the remaining 20, 16 were considered eligible for PrEP, 1/16 (6.3%) decided not to take it and 15 (93.8%) started PrEP as a combined intervention for HBV treatment and HIV prophylaxis. When investigating follow up and retention in care, out of the 14/15 (93.3%) that were due for a refill, 8/14 (57,1%) returned for at least 1 refill, amongst whom 6/12 (50%) had two or more refills (Suppl figure 1).

**Conclusion:** Sexual health and PrEP programmes provide an important opportunity for HBV testing and treatment for young adults across high HIV burden settings. However, attrition from the care cascade at each step highlights the pressing need for interventions that address barriers to sustainable delivery of long-term care. Our HBV and PrEP programmes continue working to support education, clinical evaluation and service development for HBV in these populations.

## Introduction

High profile global goals have been established for the elimination of hepatitis B virus (HBV) as a public health threat, with specific targets for the year 2030 [1]. Nucleos/tide analogue (NAs) agents are offered to those deemed at the highest risk of chronic liver disease to reduce morbidity and mortality, and to reduce transmission. Tenofovir disoproxil fumarate (TDF), the first line agent, is included in the WHO essential medications list, and should be widely accessible and affordable (global benchmark price US $2.40/month) [2]. However, the practical reality is that TDF is not available or incurs high out-of-pocket costs for many high-prevalence populations, including the World Health Organisation (WHO) African region, which now accounts for >60% of new HBV infections [2].

In March 2024, the WHO published new guidelines for HBV [3], which simplify and widen treatment eligibility criteria, and for the first time endorse dual therapy when TDF monotherapy is not available. This provides flexibility for TDF to be prescribed as part of fixed-dose combination therapy, together with either lamivudine (3TC) or emtricitabine (FTC), collectively termed ‘TDF/XTC’ [3]. In many settings, TDF/XTC is more affordable and accessible than TDF monotherapy as a result of its widespread procurement for HIV treatment. These fixed-dose combinations are also used as Pre-Exposure Prophylaxis (PrEP) in adolescents/adults who are at risk of HIV acquisition. PrEP therefore offers the combined benefit of HBV treatment and HIV prevention [4]. Such approaches may be of particular strength in many African settings, where HBV programmes can build on expertise, infrastructure and resources that have been developed for tackling HIV [5].

In South Africa (SA), HIV and HBV infection are co-endemic [6, 7]. HIV has been tackled through scale-up of education, screening and treatment, leading to proactive community engagement and robust access pathways to diagnosis and treatment. Antiretroviral therapy (ART) is free of charge through the Global Fund and the United States President’s Emergency Plan for AIDS Relief. In comparison, HBV infection has been neglected [8, 9], with poor awareness, high stigma, and low access to interventions for prevention, diagnosis and treatment [10–12].

We here describe the results of a collaboration between translational research studies for provision of HIV-PrEP and HBV care pathways, based at the Africa Health Research Institute (AHRI) in KwaZulu-Natal (KZN), SA. We set out to gather preliminary data to explore the prevalence of HBV infection among adolescents and young adults (AYA) engaging with sexual health screening and PrEP services, and to determine the uptake of PrEP (for people testing HBsAg-positive).

## Methods

### Study setting and research cohorts

We developed a collaboration between programmes at AHRI (**Figure 1**). The ‘EVOLVE-HBV’ study started in 2023 as a collaboration between AHRI and University College London (UCL) and the Francis Crick Institute in the UK, to describe HBV epidemiology in the KZN population, characterize the clinical and laboratory features of HBV infection, and engage with local communities and healthcare providers to implement sustainable care pathways to diagnosis [10, 13]. This study is approved by the University of KwaZulu-Natal (UKZN) Biomedical Research Ethics Committee (BREC) (ref. 00004495/2022) in SA, and UCL ethics committee in the UK (ref. 23221/001 EVOLVE-HBV).

**Figure 1:**
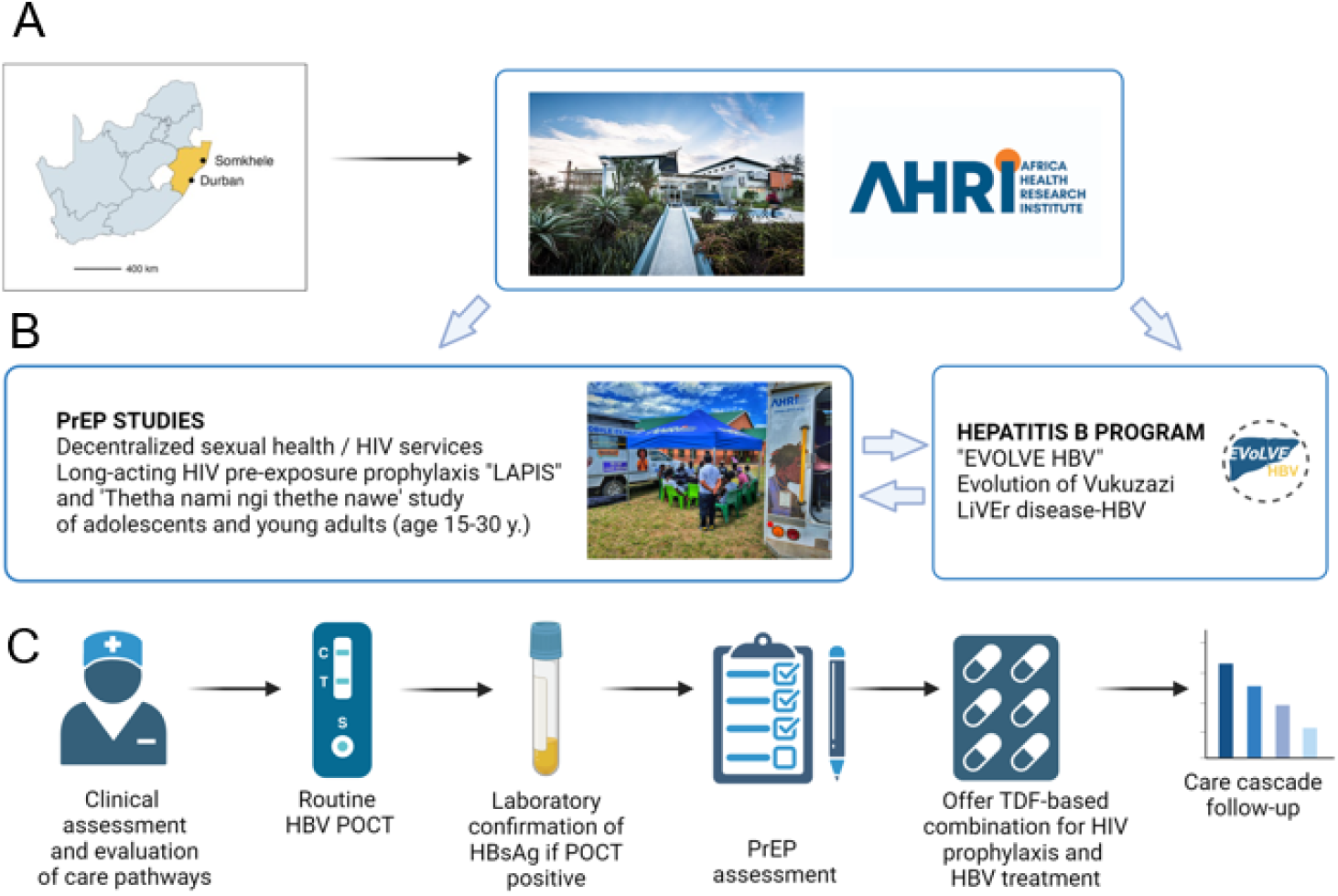
Schematic structure of studies at the Africa Health Research Institute (AHRI) to investigate frameworks for delivery of sexual health services and HIV pre-exposure prophylaxis (PrEP) as a foundation for delivering testing and treatment for HBV. A: Location of study sites; map shows South Africa with the KwaZulu-Natal province coloured gold and the two AHRI sites of Durban and Somkhele highlighted. B: Collaboration between PrEP studies and HBV program. C: Pathway offered to adolescents and young adults showing HBV screening, assessment, offer of PrEP and follow-up.

To explore the role and impact of programmes providing HIV PrEP for tackling HBV infection in AYA aged 15-30 years, we used a framework established by ‘*Thetha nami ngithethe nawe*’ (‘Talk to me and I’ll talk to you’ in IsiZulu) [14], which is a cluster-randomised study established to investigate the effectiveness, implementation and cost effectiveness of peer-led social mobilisation through decentralised HIV and sexual reproductive health (SRH) services, approved by UKZN BREC (ref. 473/2019). Young people in the community are engaged by peer navigators, and offered a needs assessment. Those at risk of sexually transmitted infection (STI) are referred for review in nurse-led mobile clinics that provide testing and treatment, offer contraception, preconception care, and tailored HIV prevention. PrEP is offered as a continuous (rather than event-based) intervention to those who are sexually active, and willing to engage with taking daily medication and attending follow-up. *Thetha nami* has also provided a platform for the long-acting HIV PrEP (*LAPIS*) study, approved by UKZN BREC (ref. 3735/2021). LAPIS is a cluster-randomised controlled trial of effectiveness and implementation of offering injectable PrEP, continuous oral daily PrEP, vaginal rings, or packs of post exposure prophylaxis.

### Clinical screening, and data collection

We retrospectively reviewed records of participants in SRH programmes between June 2022 - September 2024. Mobile study clinics follow a monthly schedule visiting different sites in the study area. Individuals who attend are offered HIV counselling and point of care testing (POCT), with immediate information and initiation of ART for those testing HIV positive. Individuals who are HIV negative undergo assessment for PrEP eligibility according to SA National PrEP/ART guidelines (**box 1**). The current preferred PrEP regimen in SA is a fixed-dose combination of oral TDF/FTC.

All individuals presenting at the clinics are also offered testing for STI with POCT for syphilis, and self-taken vaginal swabs or urine tests for gonorrhoea and chlamydia (**Supplementary methods**). If any of the STIs are positive these individuals receive treatment [15] and the program supports partner notification.

Hepatitis B screening is undertaken using POCT in *Thetha nami* and taking venous blood for laboratory testing in *LAPIS*. Confirmation tests are conducted using Abbott Architect System (Abbott Park, Illinois, USA). HBV vaccine is offered to those who screen HBV negative.

### Sexual health screening and follow-up

For individuals who screened positive for HBsAg, we reviewed their progress using the care cascade for HBV (**Figure 2**), focusing on testing (offer and uptake), referral to study physician, PrEP uptake and return for refill. Once participants have been initiated for PrEP, they are scheduled for a mobile clinic appointment one month after PrEP initiation. As per national guidelines, refills and monitoring are every 3 months thereafter through mobile clinic appointments [14]; we provide community refills aiming for continuous PrEP supplies. For those who do not return for a PrEP refill, the clinical and study teams aim to make contact by phone, peers or trackers (providing home visits).

### PrEP initiation and follow-up time definitions

- Time 0 - initiation visit (30 day PrEP supplied)
- Time 1 - First refill (90 days PrEP supplied)
- Time 2 - Second PrEP refill (90 days PrEP supplied)

## Results

### Community screening for HBV

Over the time period reviewed (May 2021 - Sept 2024), 15,847 AYA received a ‘needs assessment’ by peer navigators in the community, of whom 3481 (21.9%) were eligible for HIV prevention interventions and referred for clinical review. Of these, 3431 (98.6%) accepted HBV screening and 21 (0.6%) tested positive for HBsAg (**Table 1)**. Overall, most of the participants were female (2019/3431, 59%) living in rural areas (2712/3431, 79%) and aged 20–24 years (1295/3431, 38%).

**Table 1:** Characteristics of adolescents and young adults attending the *‘Thetha nami’/’LAPIS’* study mobile clinics in KwaZulu-Natal, classified by Hepatitis B surface antigen (HBsAg) status.

| Participant characteristic | Total cohort<br>N=3,431 | HBsAg positive<br>N=21 | HBsAg negative<br>N=3,410 | P-value<sup>1</sup> |
| --- | --- | --- | --- | --- |
| **Sex** |  |  |  | 0.465 |
| Male | 1412/3431 (41.2%) | 7/21 (33.3%) | 1405/3410 (41.2%) |  |
| Female | 2019/3431 (58.8%) | 14/21 (66.7%) | 2005/3410 (58.8%) |  |
| **Age (median, in years)** | 22 (19, 26) | 29 (25, 30) | 22 (19, 26) | **<0.001** |
| **Age groups (in years)** |  |  |  | **<0.001** |
| 15 - 19 | 1019/3431 (29.7%) | 0/21 (0.0%) | 1019/3410 (29.9%) |  |
| 20 - 24 | 1295/3431 (37.8%) | 3/21 (14.3%) | 1292/3410 (37.9%) |  |
| 25 - 30 | 1114/3431 (32.5%) | 18/21 (85.7%) | 1096/3410 (32.2%) |  |
| **Area of residence** |  |  |  | 0.053 |
| Rural | 2712/3431 (79.0%) | 13/21 (61.9%) | 2699/3410 (79.1%) |  |
| Urban/ Peri-urban | 719/3431 (21%) | 8/21 (38.1%) | 711/3410 (20.9%) |  |

<sup>1</sup> Chi-squared or exact p-value as appropriate HBsAg: Hepatitis B Virus surface antigen

### Characteristics of individuals living with HBV

Compared to the HBV-negative population, the 21 AYA testing HBsAg positive were comparable in sex (14/21, 66.7% female), and location (13/21, 61.9% rural). However, those testing HBsAg-positive were typically older than their HBsAg-negative counterparts, between 25-30 years (18/21, 85.7%), p<0.001. The proportion of common curable STIs in AYA living with HBV was no different from those who tested HBsAg-negative (7/21 vs 668/2293, 33.3% vs 29.1% p=0.7). The proportion of syphilis in HBsAg positive was no different when compared to that of the general population (1/21 vs 55/3409, 4.8 vs 1.6% p=0.3) (Suppl **Table 1**).

### Assessment for PrEP

The 21 individuals diagnosed with HBV had not previously been aware of their HBV status, but one was already receiving ART for HIV infection. The other 20 were counselled regarding starting oral PrEP, among whom 4/20 (20%) were not eligible following assessment (**Box 1)** and 16/20 (80%) were considered eligible for PrEP as an intervention for HIV prevention.

### PrEP uptake

PrEP was taken up as a combined intervention for HBV treatment and HIV prophylaxis in 15/16 (94%) of those eligible, while 1/16 (6.3%) decided not to take it. When investigating follow up and retention in care, out of the 14/15 (93.3%) that were due for a refill, 8/14 (57.1%) returned for at least 1 refill, amongst whom 6/12 (50%) had two or more refills (Suppl **figure 1**). At the time of follow-up for HBsAg positive individuals and on PrEP, the maximum followup time was 3 months.

## Discussion

This study provides a pilot assessment of HBV screening and offering PrEP as part of a program of SRH interventions for AYA in KZN, SA. WHO guidelines have been expanded with the goal of reaching 40 million more people with HBV treatment by the year 2026, and provide flexibility for use of dual therapy TDF/XTC [2]. The use of TDF/XTC as a combination therapy for HIV PrEP and HBV treatment meets mandates for decentralisation and task-sharing [3], making interventions more accessible to wider populations, and with potential cost-savings for health systems. Capitalising on existing frameworks for HIV and SRH will improve pathways for diagnosis and access to treatment.

We found a lower prevalence of active HBV infection (0.6%) compared to HBV prevalence rates previously observed in individuals receiving HIV care, and high risk individuals (sex workers (SWs), men who have sex with men (MSM) and people who inject drugs (PWID)) in whom HBV prevalence is 8.5% and 4% respectively [16, 17]. Lower rates in the general population are in keeping with the success of HBV vaccine roll-out as part of the WHO Expanded Programme on Immunization since the mid-1990’s. However, the results also reflect gaps in population HBV immunity, with some AYA living with HBV, reflecting the lack of roll out of the birth dose vaccine, and possible gaps in coverage of the routine multivalent vaccine infant vaccine schedule. Among the small number of diagnosed PLWHB, there was a high PrEP uptake but a fall-off in PrEP continuity over time.

One individual who had tested HBsAg-positive and was deemed PrEP eligible declined this intervention. The motives for not wishing to take PrEP were not investigated, but potential barriers to PrEP uptake include pill burden, stigmatization, doubting the efficacy of PrEP, size of the pill, concerns about side effects, myths and misconceptions about PrEP use, and costs of long-term access (e.g. travelling or missed time from work to collect refills) [18].

### Caveats and limitations

The *Theta Nami* and *LAPIS* studies are successfully engaging a sexually active group with high PrEP eligibility. However, to date, we have only identified a small number of individuals who started PrEP as a combined intervention for HBV treatment and HIV prophylaxis. Although all young people in the communities targeted can access the SRH clinics, our data are not fully representative of the whole community, as those who do not engage or opt out may have specific vulnerabilities putting them at risk of blood-borne virus infections.

### Longer term aims

Future aims for HBV will include investigating the community’s acceptance of screening, treatment and preventive interventions for HBV with qualitative research to determine knowledge, beliefs and experiences, and to identify and tackle barriers. There is a need for continuous evaluation of the impact of shifting HIV interventions on the HBV treatment landscape, as HIV treatment moves towards dual therapy regimens (based on dolutegravir) and long-acting injectable PrEP, neither of which currently incorporate HBV-active agents, nor have been clinically tested in people co-infected with HIV/HBV [19]. Long-term adherence to PrEP for PLWHB is important, as there are concerns about the potential for flares of HBV associated with treatment interruptions [20], but in practice serious adverse events have not been described on PrEP cessation in PLWHB [4].

PrEP is primarily used for HIV prevention but could also be expanded to individuals at risk of HBV. The integration of SRH, HIV and HBV services offers opportunities to combine education, screening, family planning, delivery of perinatal care and prevention of mother to child transmission (PMTCT) in keeping with the global ‘triple elimination agenda’ for HIV, HBV and syphilis.

## Conclusion

The framework we present here highlights opportunities to shape and integrate HBV education, prevention, diagnosis and treatment into wider SRH programs [3]. PrEP may be a valuable way for delivering effective and consistent antiviral therapy to PLWHB, especially in settings in which infrastructure for delivery of viral hepatitis care is currently limited or absent.

**Box 1: PrEP eligibility and risk assessment questions based on South African National PrEP/ART guidelines** [21]

**Eligible for PrEP**

1. HIV negative
2. Willing and able to take a pill a day?
3. Willing to return for 3 monthly follow-ups?
4. Understand that PrEP does not protect against pregnancy and STIs (contraception or condoms needed)

**Risk assessment**

1. Participant at significant risk of acquiring HIV (adolescent and young person who wants PrEP due to self-perceived risk; key population – esp. if adolescent or young: HIV negative MSM or transgender person; engage in transactional sex or sex work; person who inject drugs)
2. Are they sexually active?
3. Number of partners?
4. Condomless sex in the past 3 months?
5. UPSI with HIV positive or serostatus unknown person past 12 months?
6. Sex under the influence of drugs or alcohol?

ART: Antiretrovirals; HIV: Human Immunodeficiency Virus; MSM - men who have sex with men; PrEP - pre-exposure prophylaxis, STI - sexually transmitted infection; UPSI - unprotected sexual intercourse.

## Data Availability

All data produced in the present study are available upon reasonable request to the authors.

## SUPPLEMENTARY MATERIAL

### Supplementary methods

Storage and collection of samples for sexually transmitted infection:

Specimen collection kits are kept at ambient range (18 - 25C). These kits are in a cooler box with ice bricks when nurses are going to different collection sites. POCT (for syphilis and HBsAg) is done from a finger-prick by the nurse at the site of clinical review and recruitment.

Vaginal swab and urine samples were transported in a cooler box with ice bricks to the Somkhele laboratory. Urine is aliquoted into Xpert transport collection tubes and then stored refrigerated (2 to 8C°). Vaginal swab samples were collected into Xpert transport tubes (at the clinic) and then refrigerated overnight in the Somkhele laboratory. Both urine and swabs were shipped to Durban refrigerated; once received in Durban, these samples are frozen (−20C°) until testing.

## Supplementary tables

**Supplementary Table 1:** Relationship between hepatitis B surface antigen (HBsAg) status and sexually transmitted infections (STIs)

| STI Characteristics | Total<br>N=3,431 | HBsAg positive<br>N=21 | HBsAg negative<br>N=3,410 | P-value<sup>†</sup> |
| --- | --- | --- | --- | --- |
| **Any STI\*** |  |  |  | **0.673** |
| No | 1639/2314 (70.8%) | 14/21 (66.7%) | 1625/2293 (70.9%) |  |
| Yes | 675/2314 (29.2%) | 7/21 (33.3%) | 668/2293 (29.1%) |  |
| **Chlamydia** |  |  |  | **0.302** |
| No | 1762/2312 (76.2%) | 14/21 (66.7%) | 1748/2291 (76.3%) |  |
| Yes | 550/2312 (23.8%) | 7/21 (33.3%) | 543/2291 (23.7%) |  |
| **Gonorrhoea** |  |  |  | **0.162** |
| No | 2157/2312 (93.3%) | 18/21 (85.7%) | 2139/2291 (93.4%) |  |
| Yes | 155/2312 (6.7%) | 3/21 (14.3%) | 152/2291 (6.6%) |  |
| **Trichomonas<sup>‡</sup>** |  |  |  | **0.618** |
| No | 1408/1513 (93.1%) | 15/15 (100.0%) | 1393/1498 (93.0%) |  |
| Yes | 105/1513 (6.9%) | 0/15 (0.0%) | 105/1498 (7.0%) |  |
| **Syphilis** |  |  |  | **0.293** |
| Negative | 3374/3430 (98.4%) | 20/21 (95.2%) | 3354/3409 (98.4%) |  |
| Positive | 56/3430 (1.6%) | 1/21 (4.8%) | 55/3409 (1.6%) |  |
| **Ever treated any STI** |  |  |  | **0.609** |
| No | 115/675 (17.0%) | 0/7 (0.0%) | 115/668 (17.2%) |  |
| Yes | 560/675 (83.0%) | 7/7 (100.0%) | 553/668 (82.8%) |  |

<sup>†</sup> Chi-squared or exact p-value
\* Any STI is the combination of common curable STIs in our setting (Chlamydia, Gonorrhoea, Trichomonas and Syphilis)
<sup>‡</sup> Trichomonas is now being tested among female participants only

**Supplementary Table 2:** PrEP uptake and retention according to Hepatitis B surface antigen (HBsAg) status among adolescent and young adult participants of sexual health screening and prevention programmes in KwaZulu Natal, South Africa.

|  | **Total**<br>**N=3,431** | **HBsAg positive**<br>**N=21** | **HBsAg negative**<br>**N=3,410** | **P-value\*** |
| --- | --- | --- | --- | --- |
| **Assessed for PrEP eligibility** |  |  |  | 1.000 |
| No | 289/3431 (8.4%) | 1/21 (4.8%) | 288/3410 (8.4%) |  |
| Yes | 3142/3431 (91.6%) | 20/21 (95.2%) | 3122/3410 (991.6%) |  |
| **Ever eligible for PrEP** |  |  |  | **0.022** |
| No | 1467/3142 (46.7%) | 4/20 (20.0%) | 1463/3122 (46.9%) |  |
| Yes | 1675/3142 (53.3%) | 16/20 (80.0%) | 1659/3122 (53.1%) |  |
| **Ever started PrEP** |  |  |  | **0.008** |
| No | 638/1675 (38.1%) | 1/16 (6.2%) | 637/1659 (38.4%) |  |
| Yes | 1037/1675 (61.9%) | 15/16 (93.8%) | 1022/1659 (61.6%) |  |
| **Due for first PrEP refill** |  |  |  | 0.110 |
| No | 8/1037 (0.8%) | 1/15 (6.7%) | 7/1022 (0.7%) |  |
| Yes | 1029/1037 (99.2%) | 14/15 (93.3%) | 1015/1022 (99.3%) |  |
| **At least one PrEP refill** |  |  |  | 0.163 |
| No | 627/1029 (60.9%) | 6/14 (42.9%) | 621/1015 (61.2%) |  |
| Yes | 402/1029 (39.1%) | 8/14 (57.1%) | 394/1015 (38.8%) |  |
| **At least two PrEP refills†** |  |  |  | **0.004** |
| No | 812/991 (81.9%) | 6/12 (50.0%) | 806/979 (82.3%) |  |
| Yes | 179/991 (18.1%) | 6/12 (50.0%) | 173/979 (17.7%) |  |
| **Follow-up time (months)**<br>**(mean, min, max)** | 2.8 (0, 26) | 7.8 (0, 22) | 2.8 (0, 26) | **<0.001** |
| **Time on PrEP (months)**<br>**(mean, min, max)** | 2.0 (0, 14) | 1.7 (0, 3) | 2.0 (0, 14) | 0.384 |

HBsAg: Hepatitis B Virus surface antigen; PrEP: Pre-exposure Prophylaxis.
\* Chi-squared or exact p-value
† The denominator reflects those who were due for their second PrEP refills

**Supplementary figure 1:**
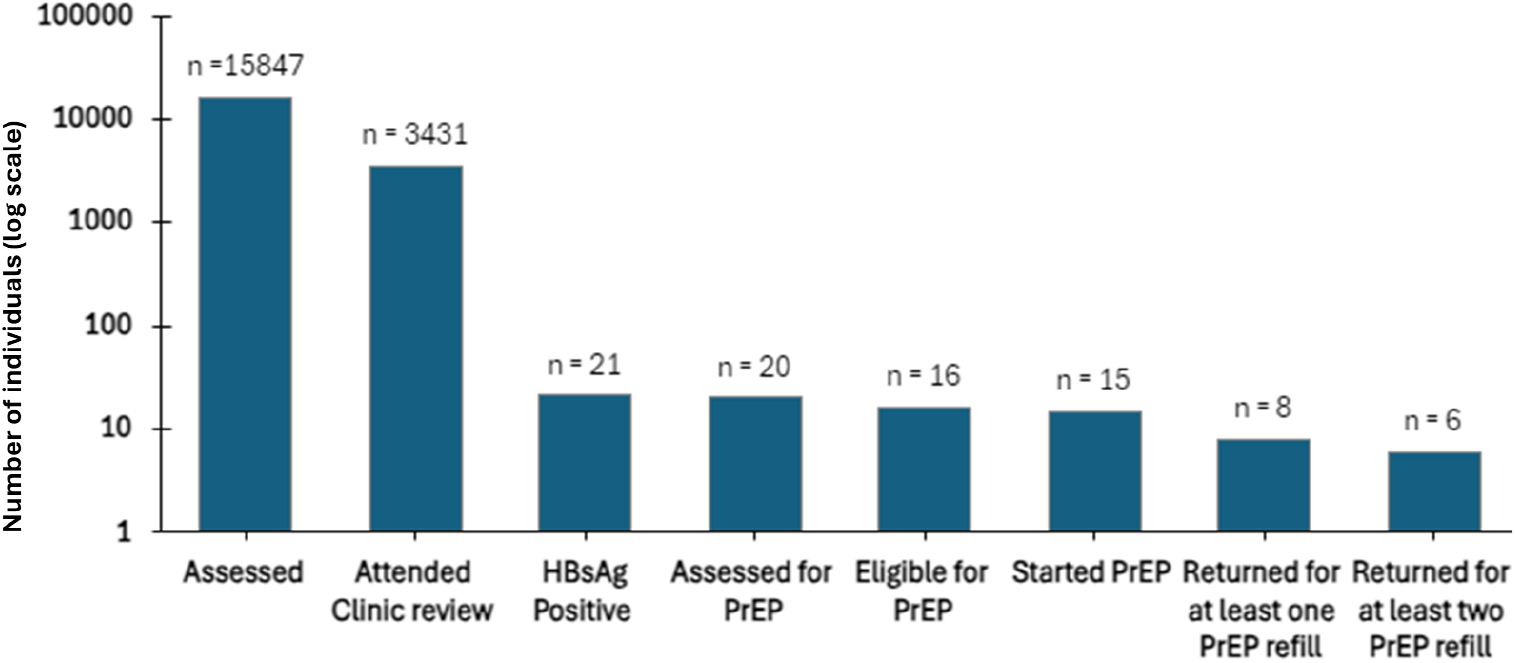
Care cascade summary of screening, linkage to care, receipt of PrEP and continuation of PrEP for young adults and adolescents accessing sexual and reproductive health services in a rural population in KwaZulu Natal, South Africa. N: number of people represented in each category of the care cascade; HBsAg: Hepatitis B surface antigen; PrEP: Pre Exposure Prophylaxis (in this case used for HIV prophylaxis and HBV treatment).

